# Comparison of Operative Time and Blood Loss with the FFX^®^ Device Versus Pedicle Screw Fixation During Surgery for Lumbar Spinal Stenosis: A Retrospective Cohort Study

**DOI:** 10.1101/2020.05.09.20096651

**Authors:** Robin Srour

## Abstract

**Objective:** Pedicle screw (PS) placement to facilitate stability and spinal fusion following lumbar decompression surgery is associated with significant soft tissue damage and blood loss. The present study evaluated differences in operative time and blood loss between PS fixation and the use of new implantable facet fusion device (FFX^®^) during surgeries of patients with lumbar spinal stenosis (LSS).

**Methods:** Patients from a single institution who underwent posterior lumbar fusion surgery for LSS with either PS fixation or the FFX device were included in this non-randomized, retrospective study. The PS group consisted of patients with LSS operated on in 2016 and the FFX group included patients operated on in 2018. Both groups excluded patients whose procedures were combined with additional implanted devices. All procedures were performed by the same surgeon. Select patient demographics, number of levels operated on, operative time and estimated operative blood loss were collected retrospectively. Differences between the two groups were assessed using the unpaired Wilcoxon two-sample test.

**Results:** Of the 70 patients undergoing fusion surgery included in the study, there were 28 in the PS arm and 42 in the FFX arm. Mean age for the PS group was 67.5 ± 9.3 years (range 42.7 to 87.5) compared to 70.4 ± 11.5 years (range 49.7 to 86.6) for the FFX group. The PS group had a greater percentage of females (57.1%) compared to the FFX group (31.0%). The mean number of levels operated were similar between the PS and FFX groups (2.3 ± 1.1 vs. 2.2 ± 1.0, respectively). Mean operative time was significantly longer for the PS group versus the FFX group (152.5 ± 39.4 vs. 99.4 ± 44.0 minutes; p<0.001). Mean operative blood loss was also significantly greater for the PS group compared to the FFX group (446.5 ± 272.0 vs. 251.0 ± 315.9 mL; p<0.001). Differences were independent of the number of levels operated on.

**Conclusion:** The use of the FFX device is associated with a significant reduction in both operative time and blood loss compared to PS fixation in LSS patients undergoing spinal fusion surgery.

## Introduction

Pedicle screw (PS) fixation following decompression is currently considered the standard technique for achieving fusion and spinal stability in patients with lumbar spinal stenosis (LSS) [1]. Placement of pedicle screws via open lumbar surgery using a posterior approach is associated with significant soft tissue damage and blood loss [2,3]. Procedural associated blood loss with PS placement increases the risk of post-operative infections, hematoma formation within the spinal canal, and blood transfusion [4,5]. The use of PS constructs can also result in adjacent level degeneration due to the rigidity produced by this approach and the resultant overload of anatomical structures [6].

The FFX^®^ device (SC Medica, Strasbourg, France) is a new implantable facet spacer designed to facilitate facet arthrodesis in patients with LSS. The device is intended to prevent facet motion and post-laminectomy instability in patients with LSS while avoiding the rigidity associated with conventional PS constructs. The FFX spacer is a titanium constructed, D-shaped device with a serrated surface which facilitates device stabilization. The device is surgically positioned between the facet joints, with its apex oriented anteriorly. Bone graft material is placed inside and posterior to the device in order to facilitate fusion.

As a result of the ability to place the FFX device under direct visualization, it theoretically reduces operative time compared to PS placement. This combined with a reduced surgical exposure requirement for the procedure versus PS potentially translates to a reduction in operative blood loss. In order to assess the above, the present retrospective cohort study was conducted with the aim of comparing differences in operative time and blood loss between procedures performed with the FFX device vs. PS in patients with LSS.

## Material and methods

The present study was a non-randomized, retrospective cohort analysis of patients with LSS undergoing spinal fusion surgery via PS fixation or the FFX device during two separate time periods. The PS group consisted of all the patients with LSS operated on in 2016 who underwent laminectomies concomitant to PS fixation. The FFX group included all the patients with LSS operated on in 2018 who underwent laminectomies concomitant to FFX fixation. These two time periods were selected as a result of PS being used as the primary implant for LSS associated surgeries during 2016 and the transition to only using the recently introduced FFX device was completed prior to 2018. Patients receiving additional spinal hardware during the same surgical admission (e.g., a combination of PS and FFX devices) were excluded from the analysis. In order to avoid the potential for operator and institutional bias, the study only included procedures performed by a single surgeon (R.S.) at the same institution. The study was approved by the local Institutional Review Board.

Data collected included patient sex and date of birth, number of spinal levels operated on, operative time and blood loss. Primary outcomes were the differences in total operative time and procedural blood loss for the PS and FFX patient populations. Secondary outcomes were the same parameters by the number of levels operated on.

### Operative Techniques

Patients in both groups were placed in the ventral decubitus or genupectoral position. A midsagittal incision was made with respect to the lumbar canal narrowness. For PS procedures, open laminectomies were performed, followed by PS fixation. Four screws were placed in the pedicles per level undergoing spinal decompression, on the right and left sides. On each side, the upper and lower screws were connected using a longitudinal rod. A transverse rod connector was used to connect right and left longitudinal rods together. Pedicle screws placement was performed using fluoroscopic guidance.

For FFX procedures, tracking of the facet joints line spacing was performed with a facet chisel followed by a reviving of the facet joints with a rasp to promote fusion. Two implants were used per level. After connecting the FFX implant onto the facet holder, bone graft material was inserted into the empty space of the device. While attached to the facet holders and at the entry of the facet joint lines, the devices were inserted into the facet joint simultaneously on the right and left sides, under direct visualization. The devices were then pushed into place using a supplied impactor and positioned appropriately. The above was followed by a laminectomy and bone graft material added posterior to the inserted implants. Surgical wounds were closed and sutured per standard routine following completion of the procedures.

### Statistical Analysis

Statistical significance for differences in surgery duration and blood loss between FFX implants and PS fixation procedures were assessed using the unpaired Wilcoxon two-sample test. All analyses were performed using R v3.6.2.

## Results

A total of 70 patients met the study criteria during the two separate time periods identified. This included 28 patients who had PS fixation in association with surgery for LSS during 2016 and 42 patients who underwent the FFX procedure for the same indication in 2018.

Patient demographics for the two study groups were similar for all parameters except for sex, with the PS group have a greater percentage of females (57.1%) compared to the FFX group (31.0%). Mean age for patients undergoing PS fixation was 67.5 ± 9.3 years (range 42.7 to 87.5) compared to 70.4 ± 11.5 years (range 49.7 to 86.6). The mean number of levels operated on were 2.3 ± 1.1 for the PS group versus 2.2 ± 1.0 for the FFX group (Table 1). The number of levels operated on ranged from one to four for both groups with each group having implants placed at various levels from L2/L3 through L5/S1. Implants were placed at L4/L5 for 24 of 28 of patients (85.7%) in the PS group and 36 of 42 patients (85.7%) in the FFX group. The mean number of implants per patient was greater in the PS group (6.5 ± 2.2, range 4 to 10) compared to the FFX group (4.38 ± 2.1, range 2 to 8) as a result of the need to place 4 screws for the initial level operated on in the PS group.

**Table 1.**
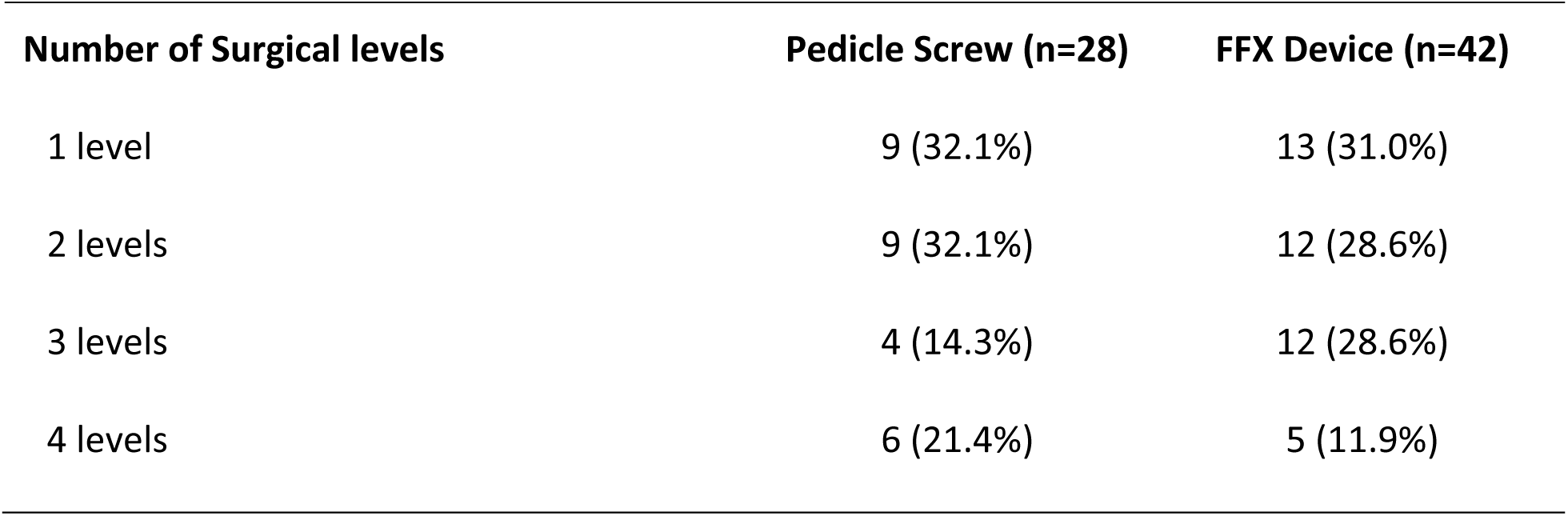
Comparison of the number of spinal levels operated on

| **Number of Surgical levels** | **Pedicle Screw (n=28)** | **FFX Device (n=42)** |
| --- | --- | --- |
| 1 level | 9 (32.1%) | 13 (31.0%) |
| 2 levels | 9 (32.1%) | 12 (28.6%) |
| 3 levels | 4 (14.3%) | 12 (28.6%) |
| 4 levels | 6 (21.4%) | 5 (11.9%) |

Mean operative time was significantly longer by 53.1 minutes for the PS group versus the FFX group (152.5 ± 39.4 vs. 99.4 ± 44.0 minutes; p<0.001) (Figure 1A). The significant difference in less operative time associated with the FFX device was independent of the number of levels operated on (Table 2).

**Figure 1.**
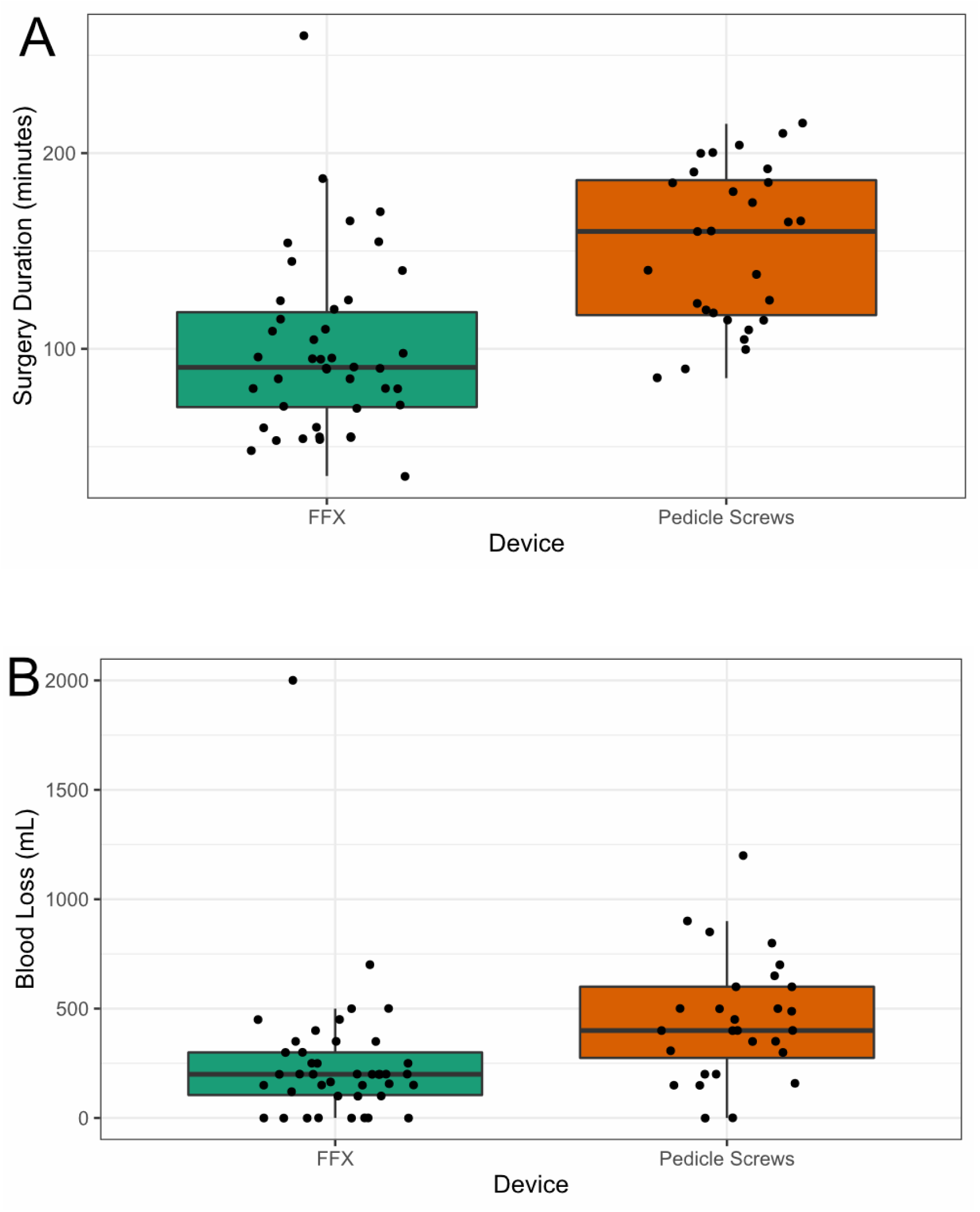
Box plots comparing A) operative time and B) blood loss for patients receiving FFX device vs. pedicle screws (PS). The difference between the FFX device vs. PS was significant (p<0.001) for both parameters.

**Table 2.** Mean operative time for pedicle screw vs. FFX device placement procedures

| **Number of Surgical levels** | **Pedicle Screw**<br>**(n=28)** | **FFX device**<br>**(n=42)** | **p value**<br>**(FFX vs. PS)** |
| --- | --- | --- | --- |
| All levels | 152.5 ± 39.4 | 99.4 ± 44.0 | p<0.001 |
| 1 level | 124.4 ± 35.3 | 90.6 ± 38.4 | p=0.03 |
| 2 levels | 133.8 ± 17.3 | 133.8 ± 38.9 | p=0.005 |
| 3 levels | 197.0 ± 15.4 | 197.0 ± 50.5 | p=0.02 |
| 4 levels | 191.0 ± 8.6 | 191.0 ± 27.5 | p=0.008 |

Operative time reported in minutes ± SD SD = standard deviation

Mean operative blood loss was also significantly greater for the PS group compared to the FFX group. Patients undergoing PS fixation experienced an average 195.5 mL greater blood loss per procedure that those in the FFX group (446.5 ± 272.0 mL for PS vs. 251.0 ± 315.9 mL for FFX; p<0.001) (Figure 1B). Mean blood loss was significantly less with the FFX device for all numbers of levels operated on with the exception of single level procedures where there was only a trend for less blood loss compared to PS fixation (Table 3).

**Table 3.**
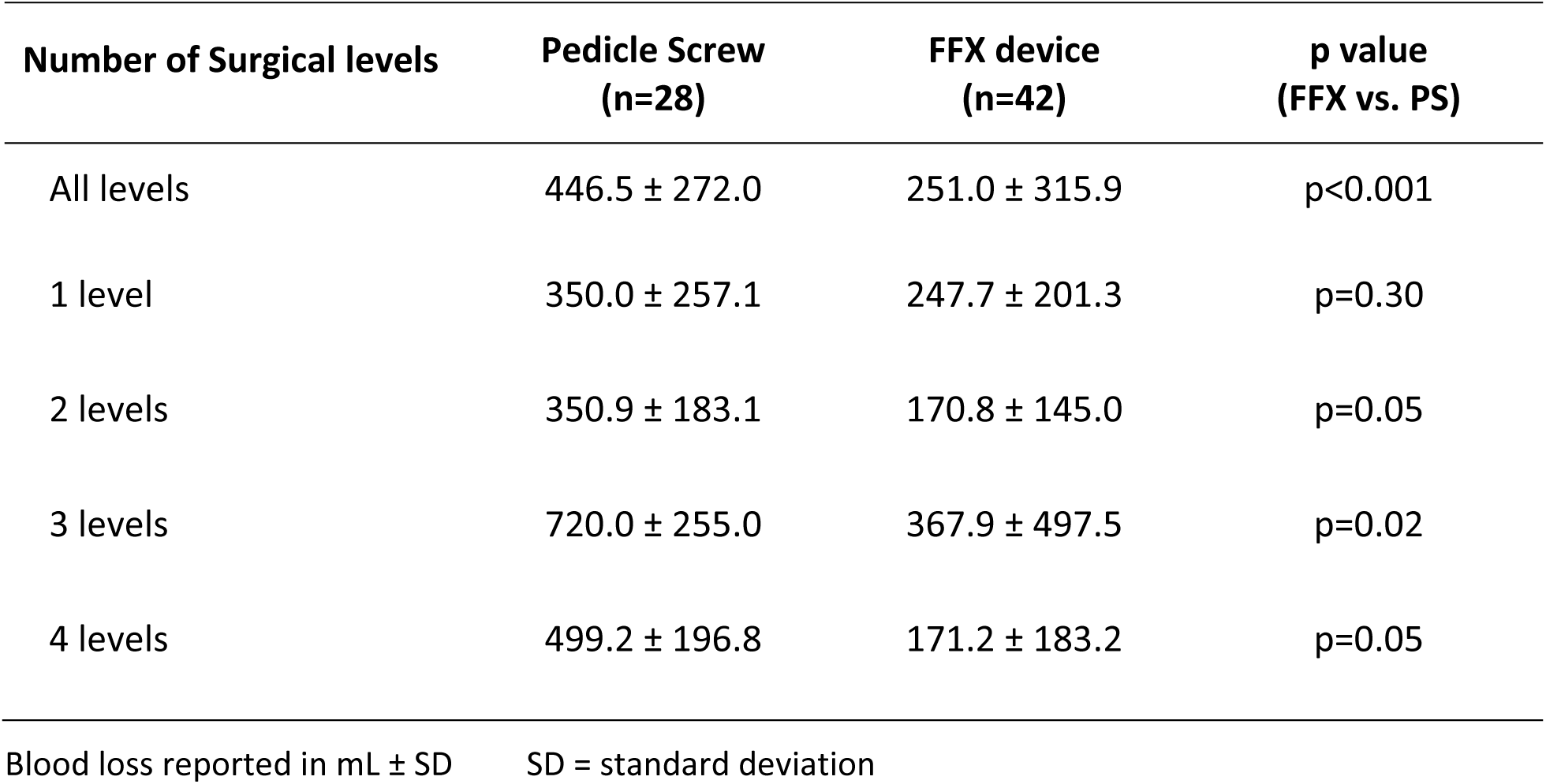
Mean operative blood loss for pedicle screw vs. FFX device placement procedures

## Discussion

The present study demonstrated that the use of the FFX device in conjunction with posterior lumbar decompression with fusion procedures for patients with LSS results in a significant reduction in both operative time and blood loss compared to PS fixation. These findings were independent of the number of levels operated.

Reducing operative time for spinal surgery procedure has several benefits. Radiation exposure is an important consideration related to the use of PS fixation for lumbar fusion since these procedures require intraoperative imaging for guidance. Cumulative exposure to ionizing radiation associated with PS placement has potential detrimental long-term effects on surgeons [7,8], with a spine surgeon’s hands and torso receiving the highest radiation doses [9]. Minimally invasive procedures to place pedicle screws have also been shown to require greater use of fluoroscopy compared to open procedure, increasing the potential for radiation-induced complications [10]. The decreased operative time and absence of the need for fluoroscopy associated with the placement of the FFX device compared to PS fixation would therefore be advantageous relative to reducing radiation exposure. Reduced operative time is also associated with decreased procedure and operating room related costs as well as enhancing operating room efficiency [11].

Increased blood loss during lumbar spine surgery is associated with the increased need for transfusions, risk of postoperative complications and length of stay following spinal surgery. Postoperative anemia has been shown to be associated with prolonged hospital stay and costs following lumbar spine procedures [12]. Additionally, receiving allogenic blood transfusions increases the risk of developing postoperative infections following spine surgery and prolonged length of stay [13]. Another concern with regard to perioperative bleeding in spinal surgery is the risk of spinal epidural hematoma formation, which might lead to spinal cord or cauda equina compression [14]. The observed reduction in blood loss with the FFX device versus PS fixation in the present study would suggest a decrease need for transfusions and associated sequalae. Additional clinical studies are needed to confirm this.

In additional to the reduced invasiveness and reduction in operative time associated with the placement of the FFX device compared to PS fixation in patients with LSS receiving laminectomies where postsurgical spinal stabilization is desired, the ability of the FFX device to provide less rigid fixation and potential reduced project loads compared to PS constructs could result in less adjacent segment degeneration and a decreased need for subsequent surgical procedures. Finite element modeling to compare the biomechanical performance of the FFX device to PS fixation both before and after fusion is obtained is needed to confirm the above as well as clinical studies which document lumbar fusion rates for the FFX device.

There are several potential limitations associated with the present study. The use of a non-randomized, retrospective study design over two separate time periods hinders the ability to make definitive conclusions when comparing these two instrumented lumbar spine procedures since confounding factors, including those which could have influence operative time and blood loss, may have impacted the outcomes observed. No attempt was made in the present study to match or stratify the two patient populations for these or other demographic factors. Additionally, the lack of detailed information regarding patient characteristics, medical history, and the criteria for the decision to operate creates the potential for selection bias. Lastly, the limitation of the patient population to procedures performed by a single individual in order to avoid potential surgeon to surgeon differences in operating time limits the ability to generalize the results. Expanding the present analysis to include additional surgeons and institutional settings would provide further evidence supporting the findings of the current study.

## Conclusion

The present study demonstrated a statistically significant reduction in both operative time and procedural blood loss associated with the FFX device compared to PS fixation in patients undergoing posterior lumbar decompression with fusion. Additional studies are needed to assess clinical outcomes with the FFX device related to pain reduction and fusion rate and to better understand differences in biomechanical performance between the FFX device and PS fixation and the potential for development of adjacent segment degeneration following lumbar decompression surgery.

## Data Availability

Data available upon request.

## Ethical Approval

The study was approved by the Institutional Review Board at Hôpitaux Civils de Colmar

## Acknowledgements

The author wishes to thank Samvida Venkatesh for performing the statistical analysis for this study.

